# Early prediction of space-occupying hemispheric infarction: Development and validation of the Malignant Edema Risk Assessment (MERA) score – Results from the PREDICT-MMI study

**DOI:** 10.1101/2025.11.21.25340775

**Authors:** Alhuda Dabbagh, Florian Welle, Janine Mielke, Max Wawrzyniak, Cordula Scherlach, Karl-Titus Hoffmann, Johann Otto Pelz

**Affiliations:** Department of Neurology, University of Leipzig Medical Center, Leipzig, Germany; Institute for Neuroradiology, University of Leipzig Medical Center, Leipzig, Germany

**Keywords:** Malignant Edema Risk Assessment (MERA), acute ischemic stroke, space-occupying hemispheric infarction, malignant brain edema, large vessel occlusion

## Abstract

**Background:** Space-occupying hemispheric infarction is a severe complication after acute ischemic stroke (AIS) due to large-vessel occlusion (LVO). As decompressive hemicraniectomy improves outcome, early identification of patients at risk is clinically crucial. We developed and validated the Malignant Edema Risk Assessment (MERA) score to predict the individual risk of space-occupying infarction after LVO.

**Methods:** Four models, each an extension of the previous one, were defined a priori: Net water uptake (estimated from the first non-contrast enhanced CT (NCCT); Model 0) + volume of intracranial cerebrospinal fluid (Model 1) + clinical data (age, localization of LVO, NIHSS at admission, successful recanalization; Model 2) + laboratory parameters, that reflected the patient’s fluid status (Model 3). The best performing model was identified based on a retrospective derivation cohort composed of patients with AIS due to LVO of the anterior circulation and first NCCT within 24 hours after symptom onset, while patients for the validation cohort were prospectively enrolled between September 2023 and February 2025.

**Results:** 155 patients with AIS and LVO were included in the derivation cohort, of whom 84 (54.2%) had a space-occupying hemispheric infarction. Model 2 performed best (MERA score ≥ 66.6%: sensitivity 75.0%, specificity 86.3%, area under the curve (AUC) of the receiver operated curve (ROC) 0.853 (0.801 – 0.918)). 156 patients with AIS (28 (17.9%) patients with a space-occupying hemispheric infarction) were enrolled in the validation study. Validating the MERA score resulted in a sensitivity of 85.7%, a specificity of 85.9%, a positive predictive value of 57.1%, and a negative predictive value of 96.5%. The AUC of the ROC amounted to 0.944 (0.909 – 0.980).

**Conclusions:** The MERA score allows an early and accurate identification of patients at high risk developing a space-occupying hemispheric infarction after AIS due to LVO.

**Clinical Trial Registration:** https://www.drks.de. Unique identifier: DRKS00033266.

**Clinical Perspective:** *What Is New?:*

- This study introduces the MERA score, a simple and semi-automatic tool that integrates early CT-based and basic clinical variables to estimate the individual risk of malignant brain edema in patients with large-vessel occlusion stroke.

*What Are the Clinical Implications?:*

- Because all input data are available before patients reach the stroke unit or neurointensive care unit, the MERA score enables early and practical risk stratification.
- An estimated cutoff of 66.6% allows clinicians to classify patients into higher-and lower-risk categories for the development of malignant brain edema using a straightforward and quickly applicable method. 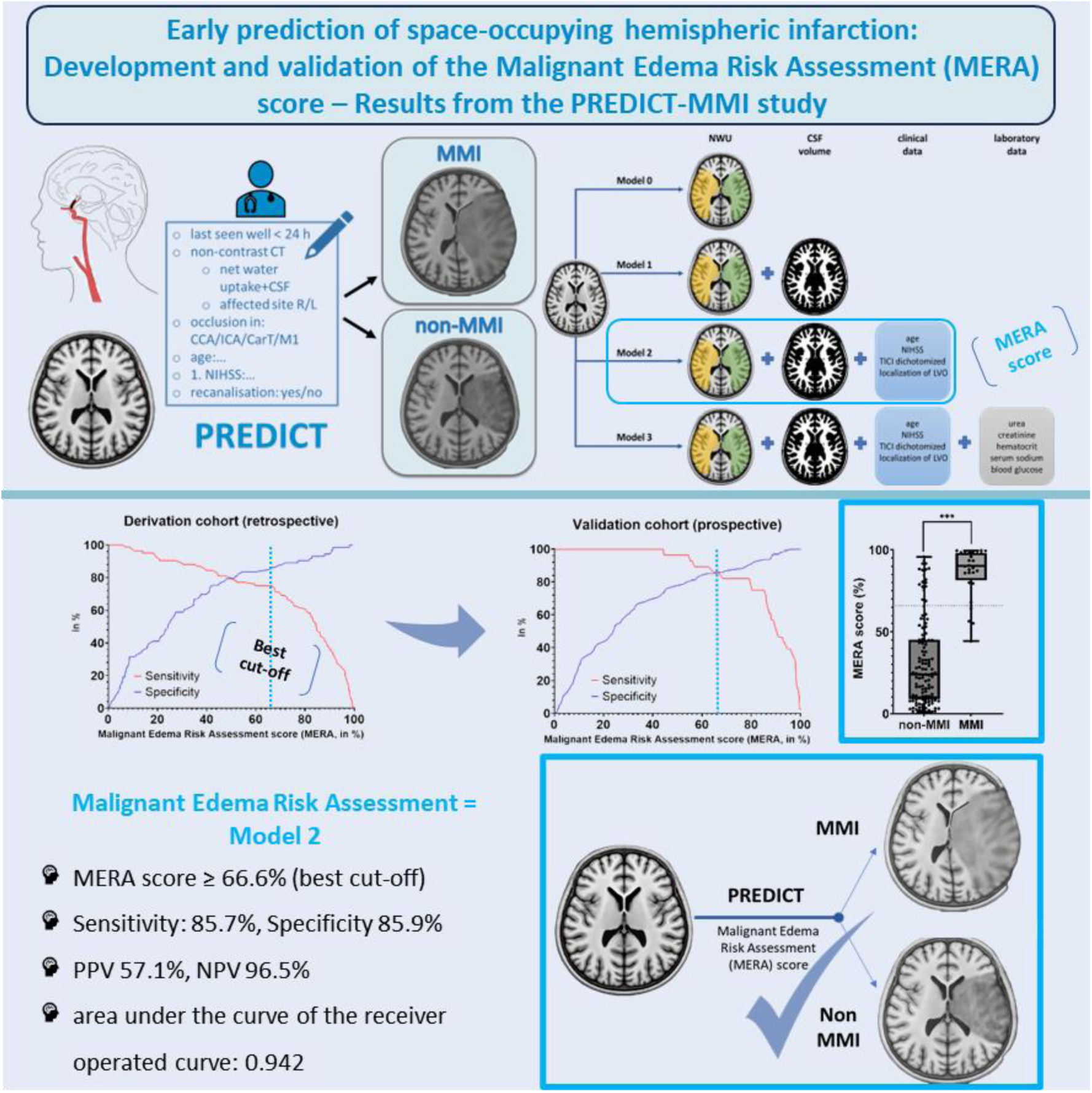

## Introduction

Space-occupying hemispheric infarction (synonymous: malignant middle cerebral artery infarction (MMI) or malignant brain edema after acute ischemic stroke (AIS)) is typically caused by large vessel occlusion (LVO) and is characterized by mortality rates of up to 80% or long-lasting disability in most survivors ^1,2^. Currently, treatment strategies focus on early decompressive surgery to provide additional space for the progressive edema to prevent fatal cerebral herniation ^3,4^.

So far, therapies that targeted the space-occupying effect of the edema like hypothermia ^5^ or glyburide ^6^ showed no benefit. One reason could have been, that patients were selected too late, i.e. after decompressive surgery ^5^ or solely based on the size of their ischemic core lesion ^7^. However, even in cases with an infarction of more than 2/3 of the territory of the middle cerebral artery (MCA), not all patients need to develop a space-occupying edema ^8^, which might have attenuated therapeutic effects in former studies. Therefore, the early identification of patients with LVO at high risk of developing a space-occupying hemispheric infarction might offer the opportunity to timely start a drug-based anti-edematous therapy, to monitor the brain edema more closely (e.g. by transcranial ultrasound ^9^), and, might finally result in a better timing of decompressive surgery.

Noteworthy, there are numerous studies, that aimed to predict the development of space-occupying hemispheric infarction in patients with LVO ^10^. However, many studies examined retrospectively collected data without a prospective validation (e.g. recently: ^11^). Moreover, study designs were very heterogeneous regarding the timepoint of risk assessment (up to 48 hours after symptom onset ^12^, the imaging modality of choice (non-contrast enhanced computed tomography (NCCT), perfusion CT, CT-angiography, magnetic resonance imaging (MRI)), or the included parameters. Finally, the risk for the development of a space-occupying hemispheric infarction was mostly calculated for groups and not on an individual basis. Sensitivity and specificity of the few clinical scores like the Malignant Brain Edema (MBE ^13^) score or the Enhanced Detection of Edema in Malignant Anterior Circulation Stroke (EDEMA ^14^) score were only moderate ^12^.

The aim of this study was to develop and validate a simple risk score, which allows to identify individual patients with a high risk for the development of space-occupying hemispheric infarction after LVO early on (i.e. already at admission to the stroke unit or neurointensive care unit).

## Methods

This non-interventional study was performed according to the ethical standards laid down in the 1964 Declaration of Helsinki and its later amendment. It was approved by the local ethics committee of the medical faculty at the University of Leipzig (reference number 240/23-ek) and registered at the German Clinical Trial Register (DRKS00033266). All participants or their legal representatives gave informed and written consent for their participation in the prospective part of the study.

### Patients

Patients with acute ischemic stroke due to LVO in the anterior circulation (internal carotid artery, carotid-T, M1 segment of the middle cerebral artery), who were treated at the Department of Neurology at the University Hospital Leipzig between January 2012 and December 2021 (*retrospective* part), respectively, between September 2023 and February 2025 (*prospective* validation) were enrolled.

Inclusion criteria for the *retrospective* part were: (1) age ≥ 18 years, (2) first ischemic stroke due to acute LVO in the anterior circulation (confirmed by CT-angiography), (3) onset of stroke symptoms less than 24 hours before the first NCCT, and (4) no intracranial bleeding (neither hemorrhagic infarction nor parenchymal hemorrhage) on the initial and first follow-up head CTs. Inclusion criteria for the *prospective* validation were: (1) age ≥ 18 years, (2) ischemic stroke due to an acute LVO in the anterior circulation, (3) onset of stroke symptoms less than 24 hours before the first NCCT. Exclusion criteria were: (1) any intracranial bleeding on the first NCCT and (2) application of radiocontrast agents before the first CT scan (e.g. during a cardiac intervention like transcatheter aortic valve implantation). Patients were excluded *during* the study, in case of: (1) death within 7 days after symptom onset before the occurrence of an endpoint (e.g. death due to change to palliative care), (2) occurrence of a space-occupying intracerebral hemorrhage within 7 days after symptom onset.

For the retrospective part, data were extracted from medical records. For each parameter, the first recorded value was used.

The primary endpoint of a space-occupying hemispheric infarction was fulfilled in the following cases: (1) the patient underwent decompressive surgery (hemicraniectomy) within 7 days due to space-occupying ischemic stroke, (2) a midline shift of ≥ 5 mm in head CT or on transcranial ultrasound within 7 days, or (3) a reduced consciousness (at least somnolence) in conjunction with a new dilated and fixed pupil on the side of infarction within 7 days (in case no cerebral imaging was performed due to palliative care).

### Models and image processing

Based on recent meta-analyses ^15,10^, and under the condition of only using routine data that was available at the time point of admission to the stroke unit / neurointensive care unit, four models for the risk prediction of a space-occupying hemispheric infarction were defined a priori (Table 1, Figure 2). We used a generalized linear model (GLM), which resembles a linear regression analysis, with a logit-link function (i.e. logistic regression) due to the binary outcome (dependent) variable.

**Table 1:**
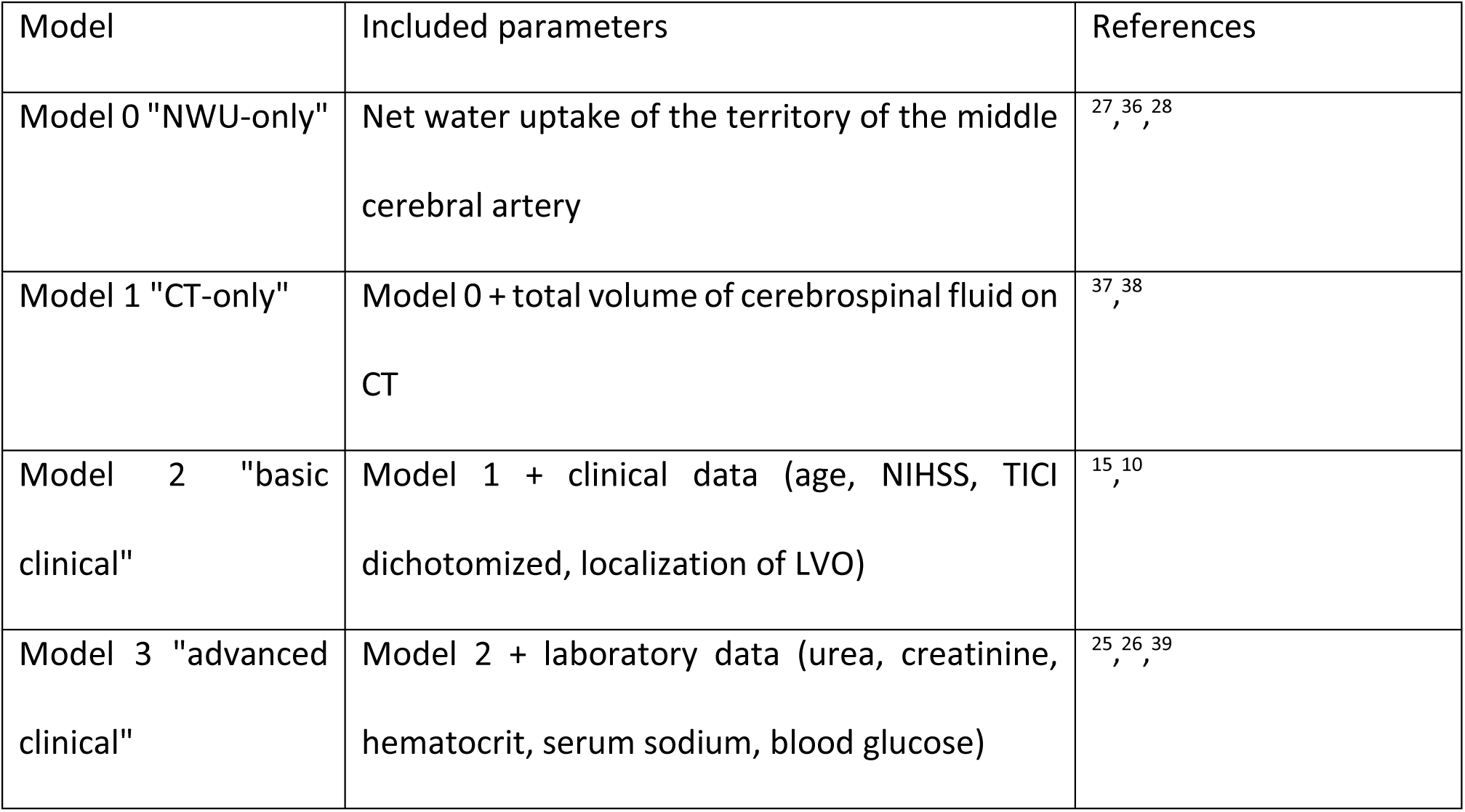
The four models, each building upon the preceding one, that were assessed for their eligibility to predict the development of a space-occupying hemispheric infarction after ischemic stroke due to large vessel occlusion (LVO). The chosen parameters were defined a priori, based on previous studies and theoretical considerations. NWU, net water uptake; CT, computed tomography; NIHSS, National Institute of Health Stroke Scale; TICI, thrombolysis in cerebral infarction.

**Figure 1:**
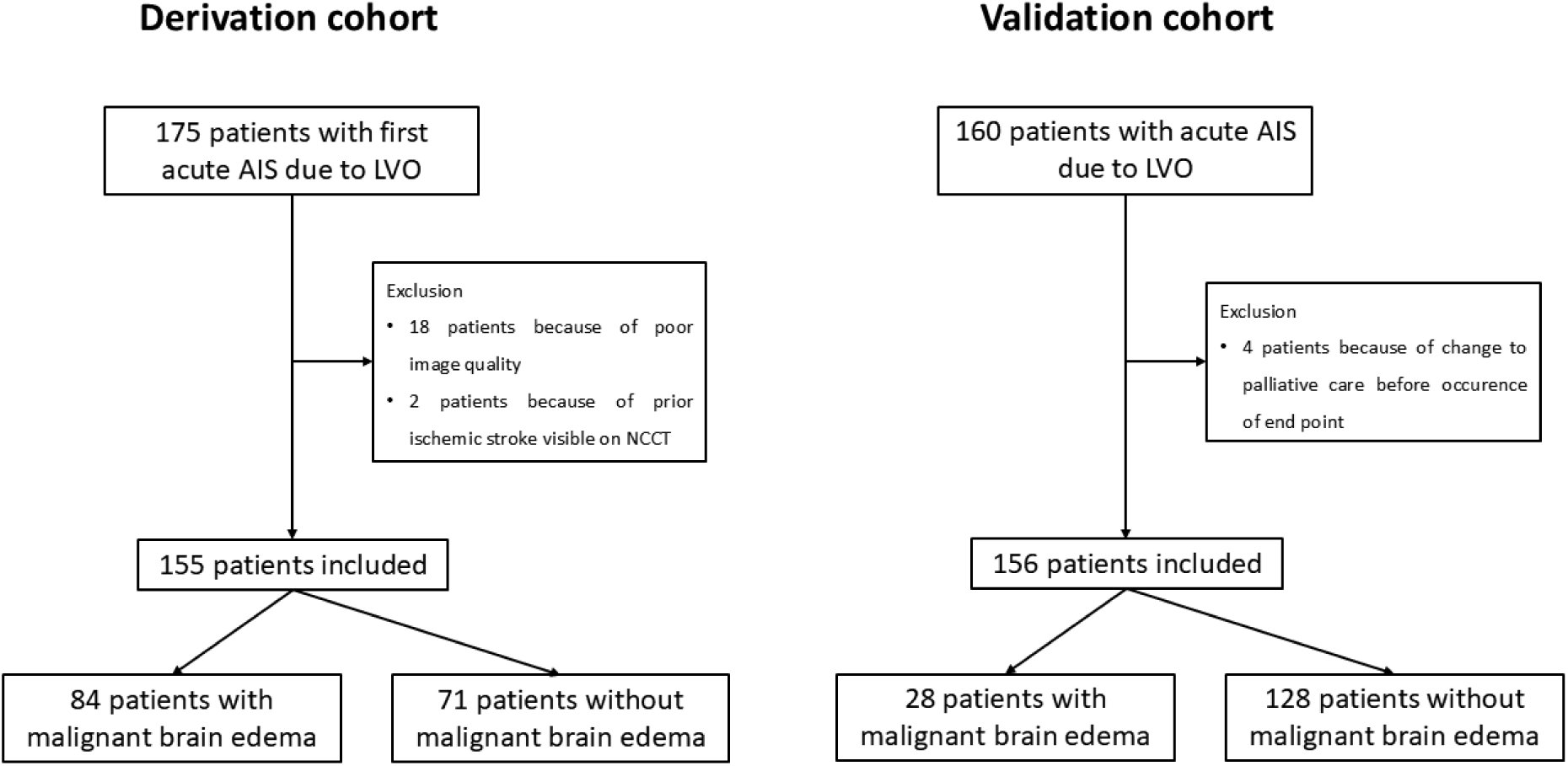
Flow chart of the study participants’ recruitment. AIS, acute ischemic stroke; LVO, large vessel occlusion; NCCT, non-contrast enhanced computed tomography.

**Figure 2:**
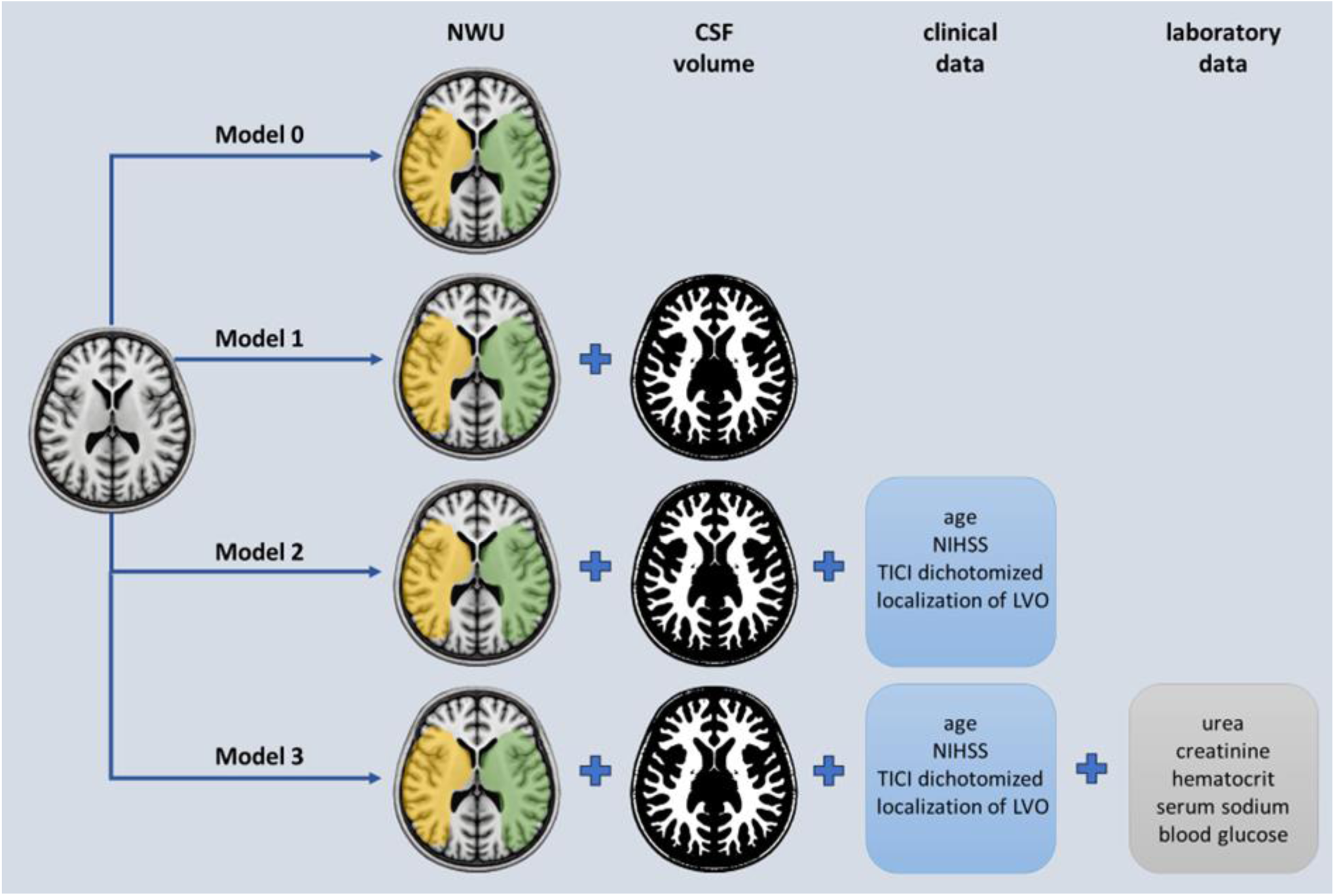
Overview over the four models for the risk prediction of a space-occupying hemispheric infarction, each building upon the preceding one, that were defined a priori. NWU, net water uptake; CSF, cerebrospinal fluid; NIHSS, National Institute of Health Stroke Scale; TICI, thrombolysis in cerebral infarction; LVO, large vessel occlusion.

Processing of the imaging data was performed using Matlab® (MathWorks®, Natick, Massachusetts, USA; Version R2023a) and consisted of the following steps:

(1) Acquisition of image data:

The first NCCT was used for further processing, either from our hospital, or in case the patient was transferred, from the referring hospital. Thus, CT scanners and protocols for acquisition of the NCCT varied in the dataset. For each patient we selected the scan with a slice thickness nearest to 5 mm and carefully checked the resulting files for quality after each processing step.

(2) Preprocessing (conversion, pseudynomization, co-registration, normalization):

The raw DICOM files were converted to the NIFTI-format ^16^ and pseudonymized ^17^. The next preprocessing steps consisted of co-registration and normalization of the imaging data to the Montreal Neuroscience Institute (MNI) space using the Clinical Toolbox ^18^ for Statistical Parametric Mapping (SPM12, Wellcome Trust Centre for Neuroimaging, United Kingdom) for Matlab®.

(3) Processing (calculation of net water uptake (NWU) and intracranial cerebrospinal fluid (CSF) volume):

The volume of intracranial CSF was calculated using the SPM12 segmentation tool. All voxels with a tissue probability of > 50 % for CSF were counted and the resulting value was converted to milliliter (ml). To account for inaccuracies of the calculated tissue probabilities in areas near the skull, the CSF mask was restricted to areas with reasonable values on the NCCT (i.e. -5 – 22 Hounsfield Units, HU). Finally, we calculated the NWU by dividing the mean HU of the MCA territory from the lesioned side by the mean HU of the contralateral MCA territory. To improve the accuracy by including only parenchymal voxels, we restricted the calculation to individual masks with values between 0 and 80 HU. Since an accurate delineation of the ischemic core area in the acute phase of a stroke is currently not attainable, we used a general mask of the MCA territory derived from the distribution of affected brain areas combined from manually delineated follow-up images of 304 stroke patients with LVO from an in-house dataset ^17^.

(4) Quality control (image quality, processing errors):

We assessed the imaging data carefully in a dedicated quality control step. Insufficient data quality, as in movement artifacts or faulty normalization, resulted in the exclusion of that patient.

### Training

In the training step, retrospectively acquired data was used to train the four logistic models with the predefined sets of predictors. We used the method of leave-one-out cross-validation to prevent an overestimation of the statistical values by overfitting the model to the provided data. Each model was trained by leaving out one patient at a time and predicting the risk of said patient by applying the model to the left-out data. The resulting predicted risks from all patients were then used in conjunction with the actual outcome to assess the performance of the model (sensitivity, specificity, area under the curve (AUC) of the receiver operated curve (ROC)). This step was repeated for all four models. The best performing model, was then finally trained on the full training dataset (i.e. not leaving one patient out).

### Validation

The model with the highest AUC in the sensitivity and specificity analysis (ROC curve) was implemented in a Matlab® program with a graphical user interface to facilitate the semi-automatic calculation of the Malignant Edema Risk Assessment (MERA) score. The MERA score indicates the probability of developing a space-occupying hemispheric infarction as a percentage ranging from 0 % (no risk) to 100 % (maximum risk). The program also gives a 95%-confidence interval, providing more insight into the certainty of the calculated risk. For the prospective cohort, the calculation of the MERA score was done around the time point of admission to the stroke unit / neurointensive care unit and took about 5 minutes per patient. To ensure blinding in the validation study, the MERA score was concealed to the treating physicians.

### Statistical analysis

SPSS (Version 29.0, IBM, Armonk, New York, USA) and Matlab® (MathWorks®, Natick, Massachusetts, USA; Version R2023a) were used for statistical calculations. After descriptive analyses, statistical significance between groups was assessed by chi square test for categorical variables and for metric variables by t-test or Mann-Whitney U-test, depending on whether the respective parameters were normally distributed or not. To determine the optimal threshold for the prediction model, the Youden Index (J = sensitivity + specificity – 1; ^19^) was calculated based on the receiver operating curve (ROC). The cutoff point corresponding to the maximum Youden Index was selected to balance sensitivity and specificity and to optimize overall diagnostic performance. The Hosmer-Lemeshow goodness-of-fit test was used to evaluate the consistency between the actual risk for space-occupying hemispheric infarction and the probability predicted by the model, which was developed in the retrospective part of this study. A p < 0.05 was considered statistically significant.

## Results

A total of 175 patients were enrolled in the derivation cohort of the study. Twenty patients had to be excluded because of insufficient quality of their first NCCT, resulting in 155 patients of whom 84 (54.2%) had a space-occupying hemispheric infarction (Figure 1). The baseline characteristics are presented in Table 2.

**Table 2:** Baseline characteristics, clinical, and laboratory data for the derivation and validation cohort. Data are stratified according to the occurrence of a malignant brain edema. * Mann-Whitney-U test, # chi square test. MMI, malignant middle cerebral artery infarction, synonymous: space-occupying hemispheric infarction; SD, standard deviation; NIHSS, National Institute of Health Stroke Scale; EVT, endovascular treatment; LVO, large vessel occlusion; CSF, cerebrospinal fluid; MCA, middle cerebral artery; TICI, thrombolysis in cerebral infarction.

|  | Derivation cohort |  |  | Prospective validation cohort |  |  | Comparison between the<br>derivation and validation cohort |  |  |
| --- | --- | --- | --- | --- | --- | --- | --- | --- | --- |
|  | All | non-MMI | MMI | All | non-MMI | MMI | All | non-MMI | MMI |
| Patients (n (%)) | 155 | 71<br>(45.8%) | 84<br>(54.2%) | 156 | 128<br>(82.1%) | 28<br>(17.9%) | - | - | - |
| Age in years<br>(mean ± SD) | 69.0 ± 12.4 | 74.7 ± 10.0 | 64.2 ± 12.3 | 74.3 ± 11.3 | 74.9 ± 11.2 | 71.9 ± 11.9 | **< 0.001\*** | 0.752* | **0.006\*** |
| Sex (female (%)) | 61<br>(39.4%) | 30<br>(42.3%) | 31<br>(36.9%) | 92<br>(59.0%) | 77<br>(60.2%) | 15<br>(53.6%) | **< 0.001#** | **0.023#** | 0.183# |
| NIHSS at admission<br>(mean ± SD) | 18.1 ± 8.1 | 16.4 ± 8.0 | 19.5 ± 8.0 | 16.5 ± 6.4 | 15.4 ± 5.6 | 21.8 ± 7.0 | 0.107* | 0.619* | 0.156* |
| Systemic<br>thrombolysis (n<br>(%)) | 69<br>(44.5%) | 34<br>(47.9%) | 35<br>(41.7%) | 63<br>(40.4%) | 60<br>(46.9%) | 3 (10.7%) | 0.534# | 0.991# | **0.006#** |
| EVT (n (%)) | 100<br>(64.5%) | 59<br>(83.1%) | 41<br>(48.8%) | 125<br>(80.1%) | 114<br>(89.1%) | 11<br>(39.3%) | **0.003<sup>#</sup>** | 0.329<sup>#</sup> | 0.512<sup>#</sup> |
| Localization of LVO |  |  |  |  |  |  |  |  |  |
| ICA | 50<br>(32.3%) | 22<br>(31.0%) | 28<br>(33.3%) | 19<br>(12.2%) | 18<br>(14.1%) | 1 (3.6%) | **< 0.001<sup>#</sup>** | **0.008<sup>#</sup>** | **0.004<sup>#</sup>** |
| Carotid-T | 40<br>(25.8%) | 9 (12.7%) | 31<br>(36.9%) | 31<br>(19.9%) | 15<br>(11.7%) | 16<br>(57.1%) | 0.266<sup>#</sup> | 0.977<sup>#</sup> | 0.097<sup>#</sup> |
| M1-MCA | 65<br>(41.9%) | 40<br>(56.3%) | 25<br>(29.8%) | 106<br>(67.9%) | 95<br>(74.2%) | 11<br>(39.3%) | **< 0.001<sup>#</sup>** | **0.015<sup>#</sup>** | 0.483<sup>#</sup> |
| TICI |  |  |  |  |  |  |  |  |  |
| ≤ 2a | 89<br>(57.4%) | 29<br>(40.8%) | 60<br>(71.4%) | 35<br>(22.4%) | 15<br>(11.7%) | 20<br>(71.4%) | **< 0.001<sup>#</sup>** | **< 0.001<sup>#</sup>** | 0.809<sup>#</sup> |
| ≥ 2b | 66<br>(42.6%) | 42<br>(59.2%) | 24<br>(28.6%) | 121<br>(77.6%) | 113<br>(88.3%) | 8 (28.6%) |  |  |  |
| Net water uptake<br>of the MCA<br>territory in %<br>(mean ± SD) | 2.16 ±<br>4.07 | 1.15 ±<br>3.78 | 3.02 ±<br>4.13 | 1.42 ±<br>5.01 | 4.3 ± 4.0 | 5.97 ±<br>6.55 | **0.005<sup>#</sup>** | 0.069<sup>#</sup> | 0.105<sup>#</sup> |
| Volume of CSF in<br>normalized brain<br>in ml<br>(mean ± SD) | 271 ± 121 | 330 ± 111 | 220 ± 106 | 218 ± 95 | 225 ± 97 | 184 ± 83 | **< 0.001<sup>#</sup>** | **< 0.001<sup>#</sup>** | 0.179<sup>#</sup> |

Calculating the optimal probability threshold based on the Youden index for the four pre-defined models, Model 2 “basic clinical” (NWU, CSF volume, clinical data, Table 1) best predicted the development of a space-occupying hemispheric infarction (best probability threshold ≥ 66.6%, sensitivity 75.0%, specificity 86.3%, accuracy 79.0%, AUC of the ROC curve 0.853 (0.801 – 0.918); Figure 3). While Model 0 “NWU only” (best probability threshold 53.2%, sensitivity 64.3%, specificity 58.9%, accuracy 60.5%, AUC of the ROC curve 0.622 (0.535 – 0.711)) and Model 1 “CT-only” (NWU and CSF volume; best probability threshold 57.4%, sensitivity 69.0%, specificity 74.0%, accuracy 68.8%, AUC of the ROC curve 0.755 (0.680 – 0.831)) were inferior to Model 2 (Figure 3). Adding laboratory parameters that were supposed to reflect the patient’s fluid status (Model 3 “advanced clinical”, Table 1) did not improve prediction (best probability threshold 54.4%, sensitivity 77.3%, specificity 82.2%, accuracy 79.0%, AUC of the ROC curve 0.857 (0.798 – 0.915); Figure 3). The Hosmer-Lemeshow goodness-of-fit test demonstrated a good fit of model 2 to the data (χ² = 5.976, p = 0.650). Thus, model 2 was implemented in a MATLAB® program with a graphical user interface to facilitate the semi-automatic calculation of the risk score.

**Figure 3:**
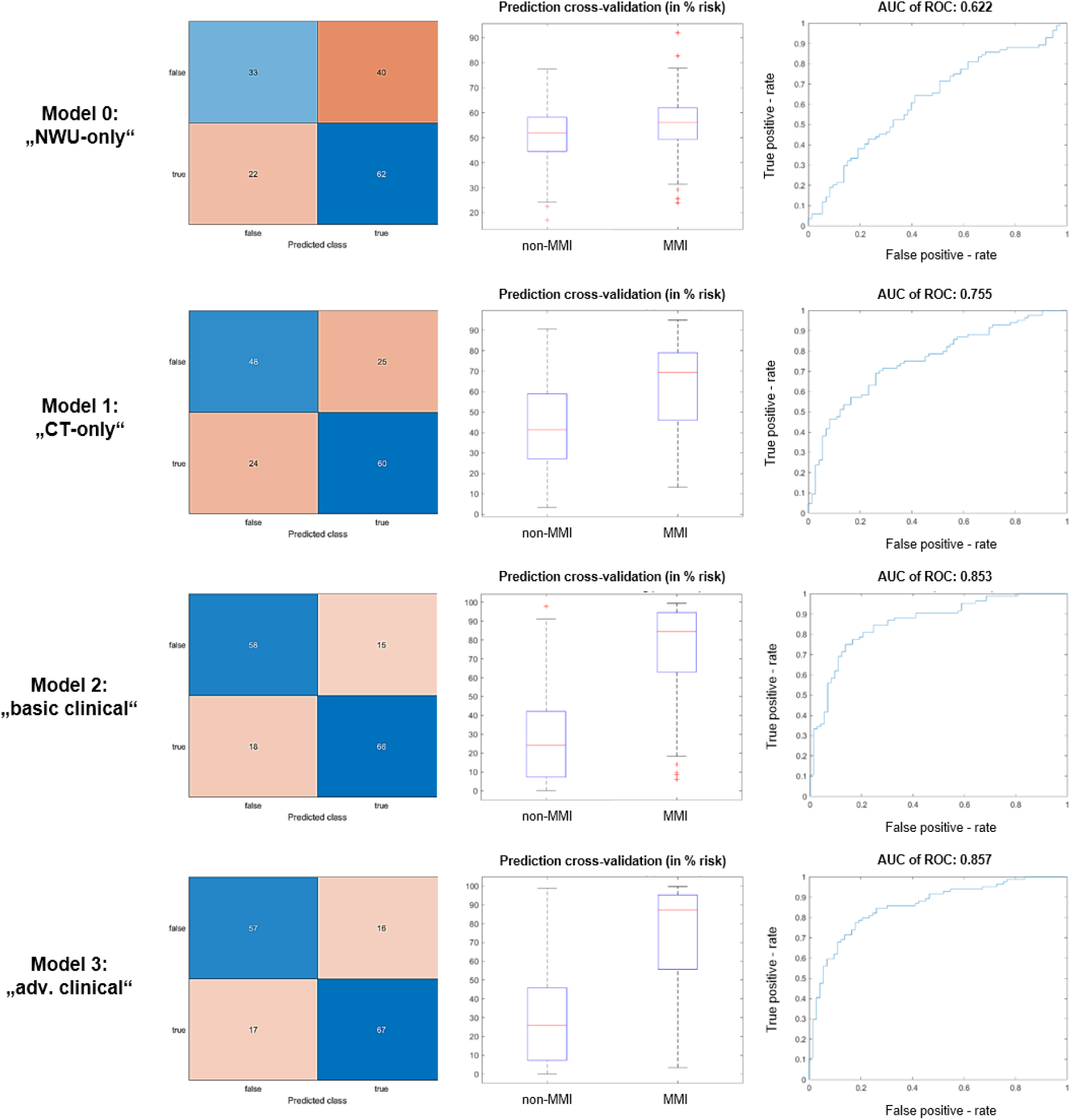
Performance of the four models in the derivation cohort. AUC, area under the curve; ROC, receiver operated curve; NWU, net water uptake; CT, computed tomography; MMI, malignant middle cerebral artery infarction / space-occupying hemispheric infarction.

Between September 2023 and February 2025, 160 patients were enrolled in the prospective validation study. Four patients were excluded because of change to palliative care and death within 7 days from symptom onset, while, at the time point of death, a primary endpoint had not occurred. This resulted in 156 patients, of whom 28 (17.9%) developed a space-occupying hemispheric infarction (Figure 1). Baseline characteristics of the prospective study cohort are given in Table 2. Compared to the derivation cohort, patients in the validation cohort were older (74.3 ± 11.3 *vs.* 69.0 ± 12.4, Mann-Whitney-U test, p < 0.001), but had a similar NIHSS score at admission (16.5 ± 6.4 *vs.* 18.1 ±8.1 Mann-Whitney-U test, 0.107). For patients with space-occupying hemispheric infarction, localization of LVO, treatment (systemic thrombolysis, EVT), and mechanical thrombectomy success (TICI ≥2b) were similar between the derivation and validation cohort (Table 2). Patients with a space-occupying hemispheric infarction had a significantly higher MERA score at admission to the neurointensive care unit / stroke unit (85.9% ± 15.8% vs. 31.3% ± 27.0%; Mann-Whitney-U test, p < 0.001; Figure 4). Using the derivation study’s MERA Score of 66.6% as threshold for the prediction of a space-occupying hemispheric infarction resulted in a sensitivity of 85.7%, a specificity of 85.9%, an accuracy of 85.9%, a positive predictive value of 57.1%, and a negative predictive value of 96.5%. The AUC of the ROC curve was 0.942 (0.909 – 0.980, p < 0.001). An overview of the sensitivity and specificity depending on the MERA score threshold is presented in Figure 5.

**Figure 4:**
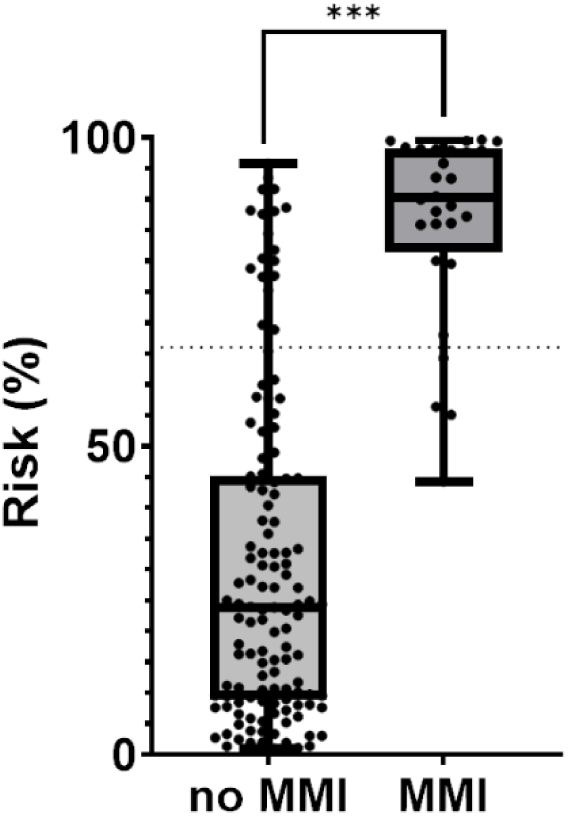
Malignant Edema Risk Assessment (MERA) scores at admission in patients with and without malignant middle cerebral artery infarction (MMI) in the validation cohort. The dotted horizontal line indicates the predefined threshold of 66.6% for risk classification. *** p < 0.001 (Mann-Whitney-U test).

**Figure 5:**
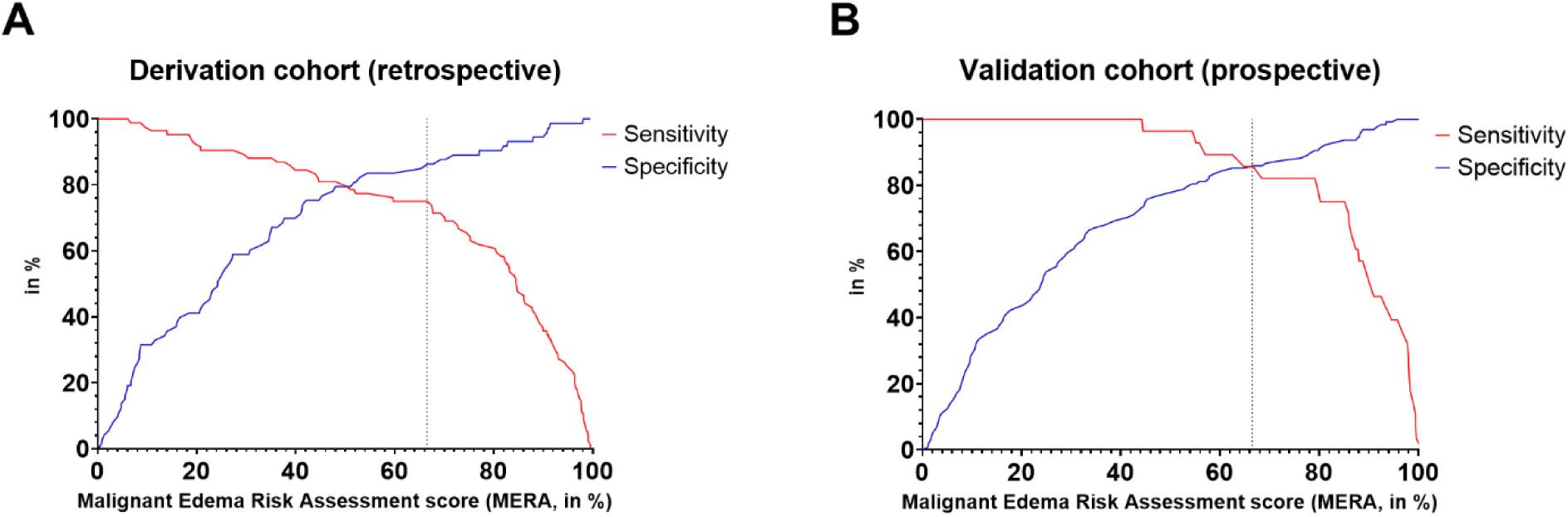
Sensitivity and specificity depending on the Malignant Edema Risk Assessment (MERA) score for the prediction of a space-occupying hemispheric infarction (derivation cohort in (**A**) and validation cohort in (**B**)). Applying the Youden index, the dotted line illustrates the MERA score (66.6 %), which allowed best prediction of space-occupying hemispheric infarction.

## Discussion

In this study, we developed and validated an algorithm (Malignant Edema Risk Assessment (MERA) score), which predicted the individual risk of developing a space-occupying hemispheric infarction after LVO with high sensitivity, specificity, and accuracy.

The introduction of EVT into the routine care for patients with acute ischemic stroke due to LVO within the last decade resulted in a significant decrease of space-occupying hemispheric infarctions ^20^. However, in patients not receiving EVT, or in those with EVT despite an already established large ischemic infarction upon presentation, space-occupying hemispheric infarction is still present with the need for decompressive craniotomy in up to 13.7% of cases ^21,22^. The early detection of patients who would likely develop a space-occupying hemispheric infarction might allow a more individualized monitoring, offer the possibility of an early anti-edematous treatment, and would be helpful for prognostication.

So far, despite promising results in pre-clinical studies, studies that aimed at reducing the malignant cerebral edema in humans with AIS were negative ^7^. One reason might have been, that patients were solely selected based on the volume of their ischemic lesion and not on the probability of developing a space-occupying hemispheric infarction ^7^. Although a large infarct volume and severe clinical symptoms were consistently associated with an increased risk for malignant cerebral edema ^15,10^, meta-analyses revealed further risk factors like younger age, a history of hypertension, a history of diabetes, or imaging parameters like a low Alberta Stroke Program Early CT Score (ASPECTS, ≤7), a low CSF volume, or an occlusion of the internal carotid *and* middle cerebral artery (carotid-T) ^10^. On the other side, there were also protective factors like smoking, successful recanalization of LVO, or lower leucocyte count at admission ^10^. Thus, patients with an infarct involving ≥ 2/3 of the MCA territory, do not necessarily have to develop a malignant cerebral edema ^8^. Vice versa, a threshold of MRI-DWI (diffusion weighted imaging) lesion volume > 82 ml predicted space-occupying hemispheric infarction with high specificity but low sensitivity ^23^. In addition, in clinical routine, time-saving multimodal CT imaging is preferred over MRI in the acute phase of stroke. Since it may be difficult to assess the ischemic core volume on early NCCT scans, hypoperfusion (in particular a reduced cerebral blood flow) on CT perfusion is considered as a surrogate for the ischemic volume. However, a recent study challenged this assumption, since the final extent of brain infarction depended on the recanalization status after EVT of LVO ^24^. Consequently, scores for the prediction of a malignant cerebral edema include several risk factors (e.g. ^25,26,12^). Therefore, we initially evaluated four combinations of well-characterized risk factors in the derivation cohort: NWU, CSF volume, age, localization of LVO, NIHSS at admission, successful recanalization (TICI ≥ 2b), and laboratory parameters that were to reflect the patient’s fluid status. Because of said reasons, we deliberately did not include predicted infarct volume into the models, as its prediction is currently still insufficiently accurate. To assure broad applicability, we focused on the initial NCCT. NWU, a kind of CT densometry, reflects the decrease of X-ray absorption of the brain tissue, which is caused by an increase of water due to an edema and / or ischemia ^27,28^. Unlike in recent studies, we did not calculate the NWU on single slices and in predefined regions of interest. Instead, we measured the NWU across the whole MCA territory, which may explain the lower NWU values in this study compared to recent studies ^27^. By adding CSF volume and established clinical parameters, the performance of the models significantly improved, which could be confirmed in the validation cohort. With the exception of the INTEP-AR score ^29^, the performance of the MERA score was better than of recently published scores (Table 3). Notably, so far, sophisticated approaches for the prediction of space-occupying hemispheric infarction like deep learning models were not superior to the use of more simple models ^30^. However, these models used hyperattenuated imaging markers, that required another NCCT, immediately ^30^ or up to 24 hours after EVT ^31^. The INTER-AR score, which was derived and validated in 2.183 patients with their first NCCT within 24 hours of symptom onset, performed better, however, it contains the occurrence of pneumonia, which is often only evident in the course of treatment, thus limiting the use of this score early after admission ^29^.

**Table 3:**
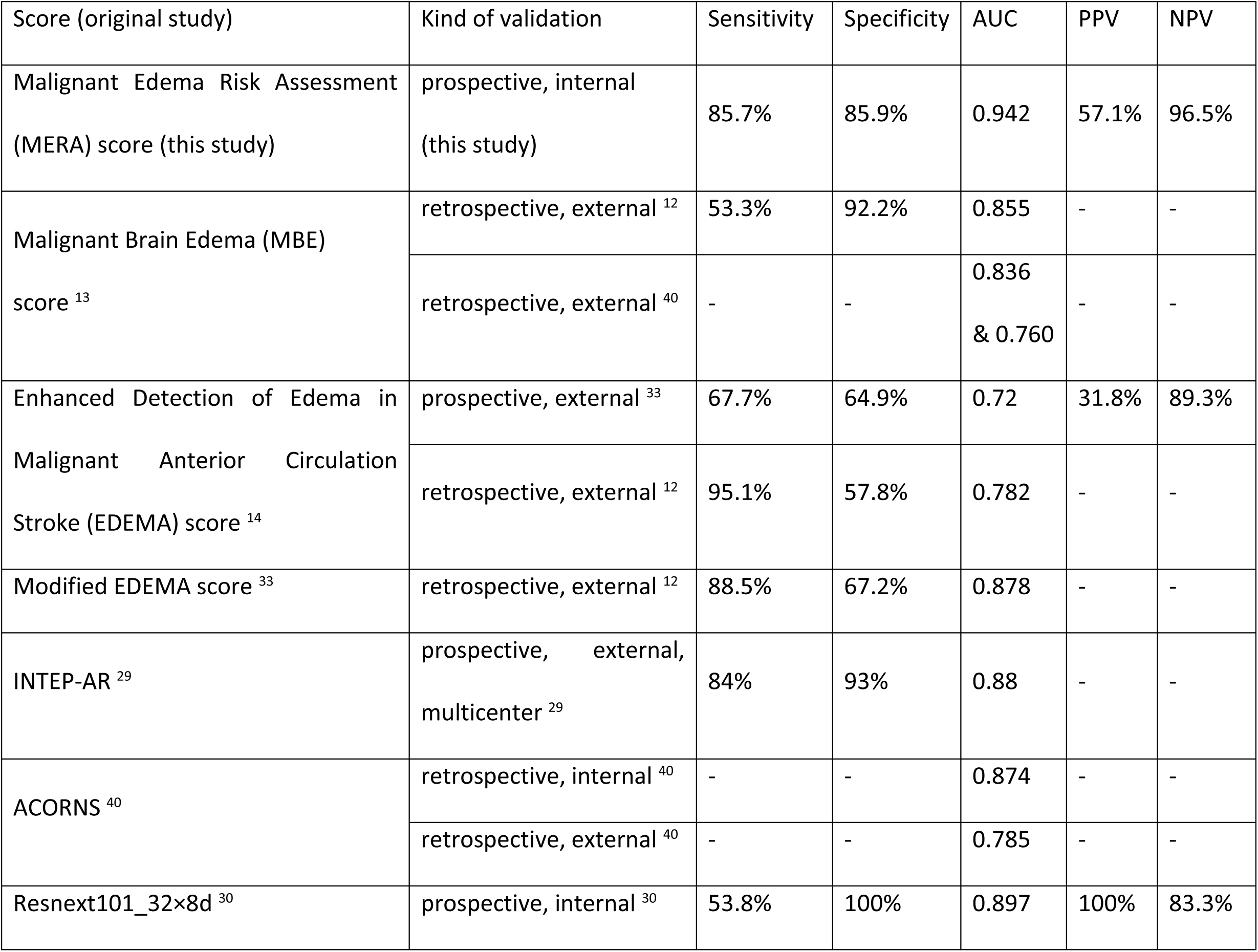
Performance of selected, validated models to predict space-occupying hemispheric infarction. Models had to be validated in an external and / or prospective cohort. AUC, area under the curve; PPV, positive predictive value; NPV, negative predictive value.

Another promising approach is the combination of transformer-based large language models with supervised ensemble learning, including static features like demographics or patient history, dynamic features like a patient’s vital signs or laboratory tests, and radiographic text to predict edema trajectory in large ischemic stroke ^32^.

Since the MERA score can be calculated even before EVT for the two recanalization states (successful or not successful recanalization), it allows the earliest identification of patients at risk for the development of a space-occupying hemispheric infarction despite EVT. Thus, anti-edematous therapies might start very early, i.e. upon admission to hospital, with the decision to continue therapy or not depending on subsequent recanalization success.

This study has some limitations. First, all patients in this study were of central European origin. So far, scores for the prediction of space-occupying hemispheric infarction were derived and mostly also validated in patients from the same ethnic origin. When the EDEMA score, originally developed in the USA, was validated on Chinese patients, specificity and sensitivity differed considerably between both populations but also between two Chinese patient cohorts ^14,33,12^ (Table 3). Thus, prospective and multi-ethnic validation studies of the different scores are needed. Secondly, the endpoint of decompressive surgery might be prone to bias. At our tertiary hospital, patients underwent decompressive surgery early (within 48 hours after symptom onset), i.e. in most cases before the full extent of the malignant brain edema was to be expected, which is in accordance with the DESTINY (Decompressive Surgery for the Treatment of Malignant Infarction of the Middle Cerebral Artery) trials ^34,35^. However, even if the NCCT before decompressive surgery did not show a relevant (≥ 5 mm) midline shift, all patients that underwent decompressive surgery had significant brain edema with edematous brain, prolapsing over the level of the skull bone on post-surgical NCCTs, thus supporting the diagnosis of a space-occupying hemispheric infarction.

## Data Availability

The data that support the findings of this study are available from the corresponding author upon reasonable request. Due to privacy and ethical restrictions related to clinical patient data, the datasets cannot be made publicly available.

## Acknowledgments

The authors declare that they have no acknowledgments to disclose.

## Sources of Funding

The authors declare that no financial support was received for the research and/or publication of this article.

## Disclosures

The authors report no disclosures or conflicts of interest.

## Notes

### Competing Interest Statement

The authors have declared no competing interest.

### Clinical Trial

Clinical Trial Registration: https://www.drks.de. Unique identifier: DRKS00033266.

### Funding Statement

The authors received no financial support from any third party for any aspect of this work.

### Author Declarations

It was approved by the local ethics committee of the medical faculty at the University of Leipzig (reference number 240/23-ek).

